# Development and Preliminary Validation of the Group Cognitive Therapy Scale to Measure Therapist Competence

**DOI:** 10.1101/2022.06.07.22276085

**Authors:** Misuzu Nakashima, Miki Matsunaga, Makoto Otani, Hironori Kuga, Daisuke Fujisawa

## Abstract

**Objective:** The aim of this research was to create a scale to assess the competency of therapists who conduct group cognitive behavioral therapy (G-CBT), which can serve as a tool to aid the continued training of therapists.

**Methods:** Three stepped studies were conducted. Study 1: Through literature review and experts’ consensus process, essential skills for G-CBT were articulated and were categorized according to the criteria of the Cognitive Therapy Scale, a well-established rating scale to evaluate clinicians’ skills in individual cognitive behavioral therapy. The list of those skills was organized into a rating scale. Study 2: Behavioral anchors were added to each skill and were classified by the levels of difficulty (beginner, intermediate, and advanced levels), based on the rating by G-CBT experts. Study 3: Interrater reliability and validity of the rating scale were examined in a sample of forty-one videotaped G-CBT sessions of actual clinical sessions and educational role-plays.

**Results:** A twelve-item Group Cognitive Therapy Scale was developed. It consists of eleven items that have been adopted from the original Cognitive Therapy Scale, with the addition of a new item called “Intervention Using Relationships with Other Participants.” This scale showed excellent internal consistency (Cronbach’s alpha: 0.95), satisfactory inter-rater reliability (interclass correlation coefficients: 0.65 - 0.88), and high predictive validity.

**Conclusion:** A novel rating scale to evaluate therapists’ competency in G-CBT was developed and successfully validated.

## Introduction

Group cognitive behavioral therapy (G-CBT) is a type of cognitive behavioral therapy (CBT) that is implemented in a group format. A group format embodies social and emotional benefits for the participants of sharing experiences with others (1). G-CBT is generally considered cost-effective compared to individual sessions (1, 2), although its effect size is somewhat lower than individual CBT. Therefore, G-CBT programs are often considered as a part of low-intensity interventions, which is provided prior to more intense interventions such as individual psychotherapy and pharmacotherapy (3). G-CBT also has been implemented outside of the medical field, for instance in the judiciary and industrial fields (4).

Although G-CBT is categorized as a low-intensity intervention, it does not mean that G-CBT is easier for therapists to conduct than individual CBT. Providing group therapy requires therapists to have skills to facilitate interactions and communication among the members of the group, to focus on the therapeutic group processes, and to find solutions to the problems that arise in the group (5, 6). Therefore, therapists of G-CBT need to have such skills in addition to the skills that are required in individual CBT.

A few manuals and competence assessment scales for group psychotherapies have been developed as the aiding tools for therapist training (7, 8, 9, 10). However, quality control methods have not been established in the field of G-CBT and awaits the development of a standardized assessment scale to define and measure therapist competence. Once developed, the scale would not only encourage self-reflection among therapists, but it would also serve as a tool for evaluating the effectiveness of training and ensuring the quality of treatment (11, 12, 13, 14).

A few scales that measure therapists’ competence in G-CBT have been developed. Hepner et al. (15) developed an adherence and competence rating scale for G-CBT for depression. The scale includes items specific to group therapy (group dynamics, group motivation, group participation, etc.). Wong (16) created a quality assessment checklist for rehabilitation group therapists of people with acquired brain injury working on their memory skills. This checklist covers competencies regarding group discussion facilitation, communication skills, interpersonal competence, and session structure.

However, these tools have limitations in the following ways: 1) they are specific to people with depression, substance dependence, or acquired brain injury and does not include skills that could apply to G-CBT for other conditions, and 2) the scales do not contain specific examples of therapist behavior, which limits the reliability of the rating. Therefore, there is a need to establish a more reliable rating scale to assess therapists’ competency for G-CBT that can be are independent of specific disorders.

The objectives of this study were to develop and validate a rating scale to evaluate therapists’ competency in conducting G-CBT. We placed emphasis on developing a scale that assesses the underlying skills common to a range of disorders and intervention methods and on developing a checklist of illustrative therapist behaviors to improve the reliability of the rating. We implemented the following processes, which are outlined in detail in the sections below. Study 1: Articulating essential skills for G-CBT by conducting literature review and obtaining experts’ consensus; Study 2: Providing behavioral anchors to each of the scale items and categorizing them by the levels of difficulty (beginner-, intermediate- and advanced-levels); and Study 3: Examining internal consistency, inter-rater reliability, and predictive validity using G-CBT video samples.

### Study 1

The objective of this first study was to articulate essential clinical skills of therapists who conduct G-CBT.

## Methods

The following three steps were taken - literature search, organizing of group therapist skills and examining face- and content-validities.

### Literature search

Articles included in the review were English publications regarding clinical competence to conduct face-to-face group therapy for adults with mental health problems. Publications regarding individual, family or online therapies were excluded. We searched peer-reviewed articles published from January 1980 to October 2020, using PsychInfo, Scopus, and PubMed databases using the broad strategy of including any of the following competence-related terms: “therapeutic factor,” “therapeutic competence,” “clinical skill,” or “clinical competence,” in combination with one or more of the following group therapy terms: “group psychotherapy,” “group format,” or “group therapy”.

A two-stage process for selecting relevant articles was employed by two independent reviewers (MM and MN). First, they screened titles and abstracts of all searched articles. Second, they reviewed full copies of screened articles and assessed for eligibility. When there was discrepancy, discussion was made until reached agreement.

### Organization of group therapist skills

To create a rating scale for G-CBT, the required skills for G-CBT which were elicited by the literature search were organized by mapping onto the framework of an existing rating scale, the Cognitive Therapy Scale (CTS). The CTS is a well-established scale to assess therapists’ competence in individual CBT. The CTS (17) and its revised edition (18) comprise eleven essential skills for CBT (agenda setting, feedback, understanding, interpersonal effectiveness, collaboration, pacing and efficient use of time, guided discovery, focusing on key cognition or behaviors, strategy for change, application of cognitive-behavioral techniques, and homework). Each item has clear goals, and a therapist’s skills are rated according to the degree of achievement of each goal on a 7-point scale (from 0 = poor to 6 = excellent), which is evaluated based on video- or audio-recordings or direct observation of actual CBT sessions. This scale has been used as the standard for therapist qualification in clinical trials and clinical practices (19, 20) and has been used in many accrediting bodies in CBT, such as the Beck Institute (https://beckinstitute.org/) and the Academy of Cognitive and Behavioral Therapies (https://www.academyofct.org/).

Four clinicians of different disciplines (a psychiatrist, a physician in psychosomatic medicine, a psychotherapist, and a clinical psychologist) with expertise in G-CBT had four focused-group discussions (six hours each) to re-categorize the extracted competencies according to the framework of the CTS. We endeavored to articulate the above essential skills in the descriptions of observable behavior. Using the CTS format, we described the essential skills under the following aspects: 1) objectives of each skill, 2) desirable therapist skills, and 3) rating criteria for each clinical skill item. Further, we added 4) a behavioral checklist for each item, so that it can serve as an objectively-measurable standard of behavior of therapists. We devised the scale so that it can be used for various mental problems and in a wide range of settings (e.g. clinical, educational, industrial, or judicial settings and stress management for healthy individuals).

We paraphrased the description in the original CTS for the group-therapy context. The descriptions in the original CTS that can be applied to G-CBT were adopted as they were, and a few new descriptions that are unique to G-CBT were added. In addition to eleven items of the original CTS, a new item “Intervention Using Relationships with Other Participants” was created, in order to accommodate skills that are specific to G-CBT. We provided each of the twelve items with 1) the objective and 2) desirable therapist strategies in group therapy. In addition, we created 3) behavioral checklists, in the scope of better illustrating the “desirable therapist strategies” as an observable behavior. Through this procedure, a prototype of the twelve-item group CTS (G-CTS) was developed.

### Face and content validities

To assess face validity of the scale, a panel of ten G-CBT experts outside the research team rated the appropriateness and importance of each G-CTS item using a five-point rating scale as follows: 1: Not important; 2: Slightly important; 3: Somewhat important; 4: Important; and 5: Very important. The members of the panel were board members of the Japanese Association of Cognitive Behavioral Group Therapy, with more than ten years of experience in G-CBT and consisted of psychiatrists, nurses, licensed mental health workers, occupational therapists, and clinical psychologists from different fields (medical, welfare, industrial, and educational).

The impact score (IS) for each item was calculated using the following formula,

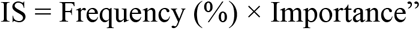

where “Frequency” is the number of experts rated the item as 4 or 5, and “Importance” is the mean score of the item. The items with the impact score of 1.5 or more were considered appropriate for the scale [21, 22].

To assess content validity, the content validity ratio (CVR) and the content validity index (CVI) was calculated.

The CVR indicates whether important and correct items are included in the scale. The CVR is calculated by the following formula;

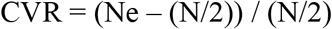

where N is the total number of experts and N is the total number of experts rated the intended item as essential. The CVR of 0.62 and more are considered appropriate [23]. The same expert panel evaluated the essentiality of the items on the following four-point scale: 4: Essential; 3: Essential but needs modification; 2: Relevant but not essential, and 1: Not essential.

The CVI was calculated to determine whether the items appropriately measure clinical skills of therapists in G-CBT. The same 10 experts rated the simplicity, relevancy, or specificity, and clarity of each item on a four-point rating scale. The number of experts who rated an item as 3 or 4 was divided by the total number of experts to calculate the CVI of that item. Items with a CVI of more than 0.90 were kept, while items with a CVI of 0.80–0.89 were revised. In addition, the items were modified based on comments in the free-text section.

Intra-class correlation coefficients and standard deviations for each value were calculated.

## Results

### Literature search

Our literature search yielded the following 148 papers after excluding duplications: thirty-eight (PubMed), seventeen (Web of Science), fifty-seven (Scopus), and thirty-six abstracts/titles (PsycInfo). Thirteen of these 148 papers were included in the review, based on the consensus review by two of our authors. Further, five relevant books were identified by manual search (24, 6, 25, 26, 27). Seventeen group therapist skills were extracted from this review (Table 1).

**Table 1.** Summary of Studies Reviewed.

| Authors, Year | Clinical Skills Required of Group Therapists |
| --- | --- |
| Gold et al., 2013 | Group climate |
| Burlingame & Barlow, 1996<sup>30</sup> | Insight |
| Schnur & Montgomery, 2010<sup>31</sup> | Therapeutic alliance, Empathy, Goal consensus/Collaboration, Group cohesion |
| Rice, 2015<sup>32</sup> | Sharing, Support, Interpersonal learning, Meaning-making |
| Gallagher, 2014<sup>33</sup> | Self-perceptions through feedback |
| Ahmed, 2010<sup>34</sup> | Catharsis, Group cohesiveness, Interpersonal learning |
| Kennard, 1987<sup>35</sup> | Acceptance, Learning from interpersonal action, Self-disclosure, Catharsis, Guidance |
| Bloch et al., 1981<sup>36</sup> | Self-disclosure, Interaction, Acceptance, Insight, Catharsis, Guidance, Altruism, Vicarious learning, Instillation of hope, Existential factor |
| Barlow et al., 2000<sup>37</sup> | Leadership to members, Interaction, Group processes |
| Joyce et al., 2011<sup>38</sup> | Altruism, Catharsis, Cohesion, Existential factors, Hope, Imitative behavior, Imparting of information, Interpersonal learning, Recapitulation of family, Socializing techniques, Universality |
| Davies et al., 2008<sup>39</sup> | Feedback |
| Brabender, 2006<sup>40</sup> | Feedback |
| American Group Psychotherapy Association, 2007<sup>24</sup> | Early establishment of therapeutic alliance, Interventions tailored to the developmental stage of the group, Understanding of subgroups, Individual member and leader roles, Executive function/Responsibility for agenda progress, Caring/Empathy, Genuineness and warmth, Emotional stimulation, Meaning-attribution, Transparency and use of self |
| Bieling et al., 2006<sup>6</sup> | Early establishment of therapeutic alliance, Individual member and leader roles, Executive function/Responsibility for agenda progress, Caring/Empathy, Genuineness and warmth, Encouragement of openness and supportive feedback between group members, Active attempt to solve any obstacles or problems within the group, Sensitivity to the stage of group development, Respect for the evolution of group dynamics and allowing the group enough autonomy for members to work with one another |
| White, 2000<sup>25</sup> | Model active participation, Model tolerance and openness to individual differences, Use of collaboration and Socratic dialogue, Communication for the universality of experiences using “we” language |
| Wagner & Ingersoll, 2013<sup>26</sup> | Individual member and leader roles |
| Lieberman et al., 1973<sup>30</sup> | Executive functions, Caring, Emotional stimulation, and Meaning-attribution. |

### Organization of group therapist skills

Table 2 shows the projection of extracted seventeen therapeutic skills onto the items of the CTS. A new twelfth item of “Intervention Using Relationships with Other Participants” was created in order to accommodate skills that are specific to G-CBT. The creation of this item was also suggested by the panel of experts who evaluated face and content validity of the scale. This item represents therapists’ skill of being aware of and utilizing the interactions of the participants (group dynamics) as a therapeutic technique. For example, a therapist may deliberately ask other participants to comment on their fellow participants, as a way of bringing insight to a single participant. Table 3 presents an example of an item of G-CTS.

**Table 2.** Relationship between Clinical Skills Required of Group Therapy Therapists and G-CTS.

| Clinical Skills Required of Group Therapy Therapists | G-CTS Item |
| --- | --- |
| Early establishment of therapeutic alliance <sup>24, 31</sup> | c, d, e |
| Interventions tailored to the developmental stage of the group <sup>24,30, 37</sup> | a, b, c, l |
| Understanding of demarcation and subgroups <sup>24, 32, 34, 37</sup> | c, l |
| Individual member and leader roles <sup>24, 26</sup> | c, d |
| Retention of executive function/Responsibility for agenda progress <sup>24, 6, 37, 27</sup> | a, d, f |
| Caring/Empathy, genuineness, and warmth <sup>24, 6, 31, 34, 35, 36, 38</sup> | c, d, e, g, h, i, j |
| Emotional stimulation <sup>24, 31, 34, 35, 36, 38, 27</sup> | b, c, d |
| Meaning-attribution <sup>24, 32, 36, 38, 27</sup> | g, h, i, j |
| Transparency and use of self <sup>24, 36</sup> | c, d |
| Utilization of the active participation model <sup>25, 36</sup> | d |
| Tolerance of and openness to individual differences <sup>25, 36</sup> | a, c, d, e, f |
| Use of collaboration and Socratic dialogue <sup>6, 25, 30, 31</sup> | e, g, h, i, j, k |
| Communication using language such as “we” and “us” to relate the universality of experiences <sup>25, 36</sup> | a, d, l |
| Sensitivity regarding group process factors and observation of important connections among members <sup>6, 34, 36</sup> | c, h, l |
| Encouragement for open-mindedness and supportive feedback among group members <sup>6, 32, 33, 39, 40</sup> | b, c, d, e |
| Active effort to solve any obstacles or problems within the group <sup>6, 32</sup> | a, c, d, l |
| Sensitivity for group development stages, respect for the evolution of group dynamics and for the autonomy of the members to work with one another <sup>6, 32, 34, 36, 37, 38</sup> | d, e, l |

<sup>a</sup>Agenda setting, <sup>b</sup>Feedback, <sup>c</sup>Understanding, <sup>d</sup>Interpersonal effectiveness, <sup>e</sup>Collaboration, <sup>f</sup>Pacing and efficient use of time, <sup>g</sup>Guided discovery, <sup>h</sup>Focusing on key cognition or behaviors, <sup>i</sup>Strategy of change, <sup>j</sup>Application of cognitive-behavioral techniques, <sup>k</sup>Homework, <sup>l</sup>Intervention using relationships with other participants

**Table 3.**
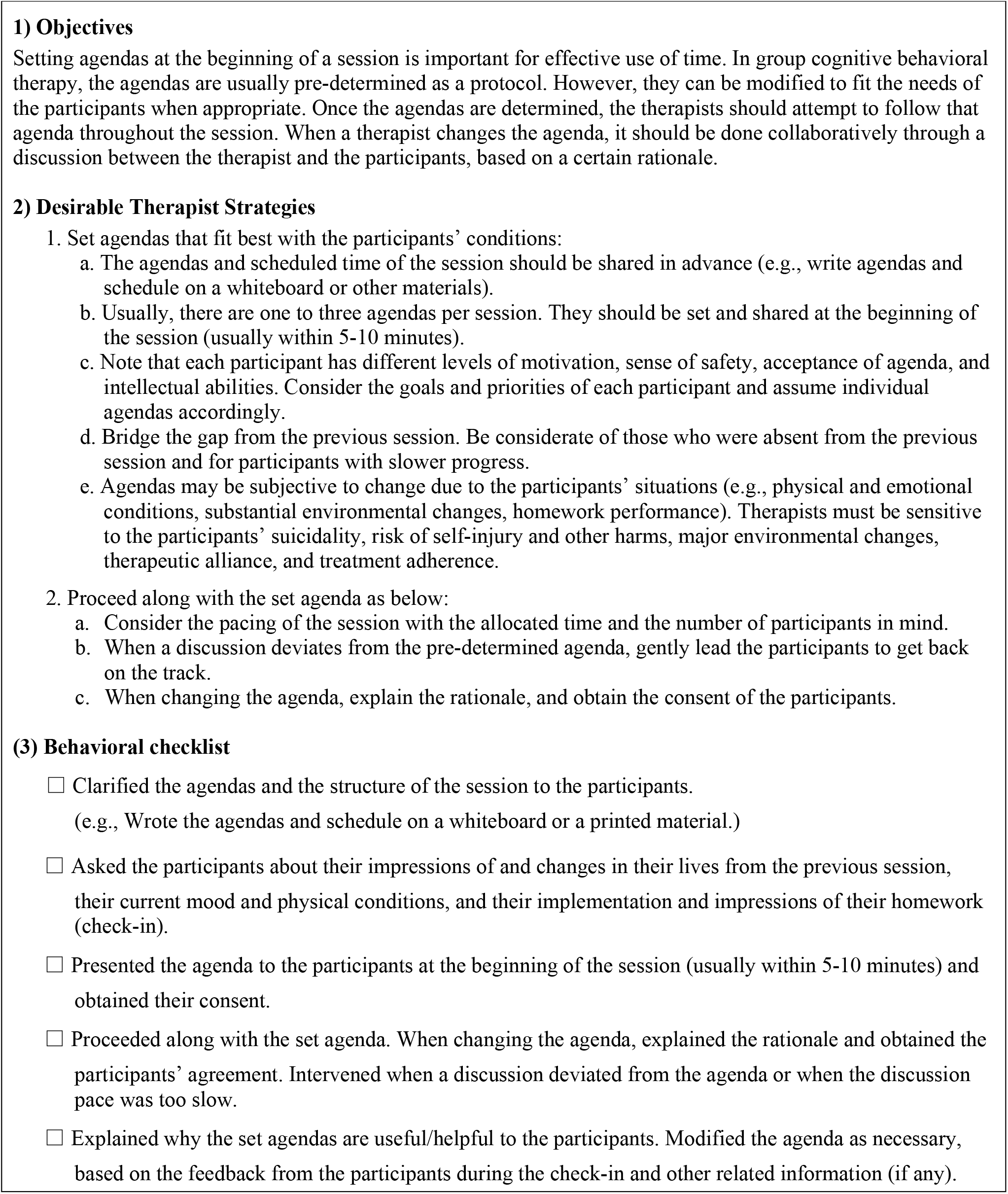
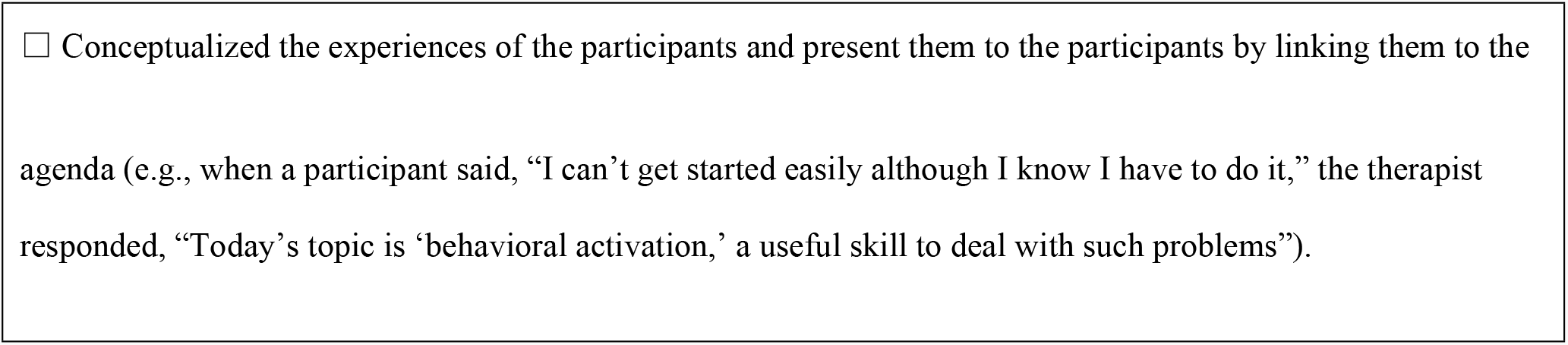
An Example of G-CTS Description (Agenda Setting)

### Face and content Validity

The Tables 4–7 shows the IS, CVR and CVI for each item of the prototype G-CTS. All of the items had the IS of 1.5 or more, thus were kept remained in the scale. The mean ICC of CVR was .719 (95° confidence interval: .607-.809), which showed a substantial agreement rate. Three items of the with the CVR of less than 0.62 and six items with the CVI of 0.80−0.89 were revised. Nine items were revised based on narrative comments by the panel. The details of the revision is described in Tables 4-7. Finally, the G-CTS with 66 behavioral-checklist items were created.

**Table 4.** Impact Score, CVR, CVI of G-CTS Items.

|  | Trial Items |  |  |  | Revision Method | Revised Items |
| --- | --- | --- | --- | --- | --- | --- |
| No | Therapist behavior | IS | CVR | CVI |  | No |
| **Agenda setting** |  |  |  |  |  |  |
| 1 | Clarified the agendas and the structure of the session to the participants (e.g., wrote the agendas and schedule on a whiteboard or printed material). | 3.36 | 1.00 | 0.90 |  | 1 |
| 2 | Asked the participants about their impressions of and changes in their lives from the previous session, their current mood and physical conditions, and their implementation and impressions of their homework (check-in). | 4.70 | 1.00 | 1.00 |  | 2 |
| 3 | Presented the agenda to the participants at the beginning of the session (usually within 5-10 minutes) and obtained their consent based on the feedback from the participants during the check-in and other related information (if any). | 3.44 | 0.60 | 0.80 | Split and partially merged with 17 | 3<br>17 |
| 4 | Proceeded along with the set agenda. When changing the agenda, explained the rationale and obtain the participants' agreement. Intervened when a discussion deviates from the agenda or when the discussion pace was too slow. Modified the agenda as necessary, based on the feedback from the participants during the check-in and other related information (if any). | 4.70 | 1.00 | 1.00 | Split | 4<br>5 |
| 5 | Conceptualized the experiences of the participants and presented these to the participants by linking them to the agenda (e.g., when a participant said, "I can't get started easily although I know I have to do it," the therapist said, "Today's topic is 'behavioral activation,' a useful skill to deal with such problems"). | 4.05 | 1.00 | 1.00 |  | 6 |
| **Feedback** |  |  |  |  |  |  |
| 6 | Checked participants' understanding and satisfaction throughout the session. | 4.05 | 1.00 | 1.00 |  | 9 |
| 7 | Checked participants' understanding and satisfaction at the end of the session. | 3.96 | 1.00 | 1.00 |  | 7 |
| 8 | Created an atmosphere that allowed the participants to express their moods and thoughts frankly, even if they were negative ones (e.g., asked a participant who remained silent during feedback and told them that even negative comments, if any, would be helpful for the group members to deepen their understanding; used self-disclosure techniques, such as asking, "The pacing of the session may have been a little too fast today. Did you find it difficult to follow?"). | 3.44 | 1.00 | 0.90 |  | 10 |
| 9 | Encouraged each participant to talk about their experiences in their own words (e.g., asked clarifying questions when a participant's remark was too abstract). | 4.60 | 1.00 | 1.00 |  | 12 |
| 10 | Received feedback from participants and shared with the participants what the therapist understood. | 3.96 | 1.00 | 0.90 |  | 8 |
| 11 | Used the feedback from the participants to conceptualize the participants or modify the behavior of the therapist as needed. (e.g., 1: Slowed down when the session pace was perceived by the participants as being too fast. 2: In response to a specific participant's comment, asked the whole group for comments in order to generalize that person's comments, such as, "Thank you for your precious comment. It must have been a bit hard for you to share it. Are there any other people who have similar opinions?") | 3.28 | 1.00 | 1.00 |  | 11 |
| **Understanding** |  |  |  |  |  |  |
| 12 | Listened to each participant and understood their thoughts and emotions accurately. | 4.60 | 1.00 | 0.90 | Split | 13<br>14 |
| 13 | Elicited remarks from each participant, understood their cognition, behavior, and emotions, and conveyed the therapist's understanding to the participants (case conceptualization). | 4.40 | 1.00 | 1.00 |  | 17 |
| 14 | Tried to understand the participants' moods through verbal and non-verbal communication and to convey the therapist's understandings through his/her attitudes and behaviors (understanding). | 1.75 | 0.80 | 0.80 | Merged with 16 | 16 |
| 15 | Understood the dynamics and processes taking place in the group and shared them with the co-leader(s) (e.g., paid attention to the occurrence of subgroups and fixed the roles of the participants). | 3.28 | 1.00 | 1.00 |  | 15 |
| 16 | Understood the participants' thoughts and feelings, not only from the information in the group session, but also from background information such as their medical history, family status, and remarks at prior sessions, and shared these with the participants (case conceptualization). | 3.20 | 0.80 | 0.80 | Merged with 14 | 16 |

**Table 5.** Impact Score, CVR, CVI of G-CTS Items.

|  | Trial Items |  |  |  | Revision Method | Revised Items |
| --- | --- | --- | --- | --- | --- | --- |
| No | Therapist behavior | IS | CVR | CVI |  | No |
| Interpersonal effectiveness |  |  |  |  |  |  |
| 17 | Was fair and honest with all participants. Didn't take condescending or deliberately humble attitudes. Did not evade participants' questions. Conveyed warmth and interest to each participant not only by the contents of his/her remarks but also with nonverbal behavior such as tone of voice and eye contact. | 4.70 | 1.00 | 1.00 |  | 22 |
| 18 | Was aware of the therapist's own emotions during the session and verbalized or used them to understand the participants' feelings. Provided appropriate self-disclosure as necessary. | 3.96 | 1.00 | 0.90 |  | 18 |
| 19 | Showed confidence that the therapist was capable of helping the participants (e.g., showed expertise related to the program or illustrated examples of thoughts and feelings that are common to many of the participants). Was not distant towards the participants. Did not show cold or intimidating attitudes. | 4.40 | 0.80 | 0.90 |  | 21 |
| 20 | Made the group a safe and secure place by balancing attending to each participant with managing the group as a whole. | 4.60 | 1.00 | 1.00 |  | 20 |
| 21 | Appropriately intervened with participants' behavior that might have hindered other participants' sense of security (e.g., competition, aggression, and imposition of opinions on others). | 3.28 | 1.00 | 1.00 |  | 19 |
| Collaboration |  |  |  |  |  |  |
| 22 | Informed the participants that their active participation was essential for the progress of the therapy (e.g., addressed a participant's reluctance to take part in the tasks in the session or homework). | 3.78 | 1.00 | 1.00 |  | 23 |
| 23 | Explained the rationale for how the program can be helpful for each participant's treatment goals. | 3.36 | 1.00 | 1.00 |  | 24 |
| 24 | Considered sharing the contents of a participant's remarks with the others so that the input could be treated as common issues in the entire group. Did not end with a 1:1 conversation between therapist and participant | 4.40 | 1.00 | 1.00 |  | 25 |
| 25 | In response to the remarks by one participant, asked other participants or the co-leader whether they had similar experiences and thereby elicited empathy and opinions. | 3.69 | 1.00 | 1.00 | Split | 26 |
| 26 | Facilitated the session by cooperatively sharing roles among the therapists – i.e., moderator, follower, confrontational. | 4.90 | 1.00 | 1.00 | Merged | 27 |
| 27 | When one aspect of the leader or co-leader's opinion is likely to have a big impact on the group, other staff members or the leader will respond to give participants a different perspective. | 4.50 | 0.80 | 0.80 | Merged with 26 | 27 |
| Pacing and efficient use of time |  |  |  |  |  |  |
| 28 | Informed the participants that the time of the sessions should be shared equally among the participants. Clarified as a rule that one participant should not speak too long. | 4.05 | 1.00 | 1.00 |  | 28 |
| 29 | Presented a rough time allocation for each agenda. | 3.96 | 1.00 | 1.00 |  | 29 |
| 30 | Progressed the session according to the pre-planned schedule. | 3.44 | 1.00 | 0.90 |  | 30 |
| 31 | Focused on important topics. When the group was distracted from the central topics and the discussion became unproductive, gently interrupted the discussion and led the discussion back to the original topic. | 4.60 | 1.00 | 1.00 |  | 31 |
| 32 | Made the discussion fruitful. When important treatment issues were not sufficiently discussed (e.g., a participant remained silent or was dedicated to unproductive chatting), the therapist cast questions to facilitate deeper discussion or explored the reason the participant(s) was not facing his/her own problems. | 3.96 | 1.00 | 0.90 |  | 32 |
| 33 | Encouraged participants to use their time equally. When a participant spent too much time on him/herself, conceptualized the participant and limited his/her talk (e.g., gently interrupted him/her by providing a summary). | 3.28 | 1.00 | 1.00 |  | 33 |
| 34 | Adjusted the session pace according to the characteristics of the participants. For hurried participants, instructed them to pace down and listen more carefully to other participants. Conceptualized and attended to the participants who were not motivated enough or who had difficulty in understanding the program. | 3.20 | 1.00 | 1.00 |  | 34 |
| 35 | Divide roles between staff members, such as facilitators and followers, confronters, etc., and work together to facilitate the group. | 4.60 | 1.00 | 1.00 | Merged with 27 | 27 |

IS: Impact Score, CVR: Content Validity Ratio, CVI: Content Validity Index

**Table 6.** Impact Score, CVR, CVI of G-CTS Items.

|  | Trial Items |  |  |  | Revision Method | Revised Items |
| --- | --- | --- | --- | --- | --- | --- |
| No | Therapist behavior | IS | CVR | CVI |  | No |
| **Guided discovery** |  |  |  |  |  |  |
| 36 | To help participants themselves become aware of their own cognitive and behavioural tendencies, first elicit detailed recollections of the situation - when, where, with whom and what they were doing - and ask about the thoughts, images that were floating and their behaviours around at the time. | 4.50 | 0.80 | 0.90 | Split and partially merged with 38 | 35<br>38 |
| 37 | Did not confront the participants. Avoided pressing or confronting issues that they were not fully aware of or were denying. | 4.30 | 1.00 | 0.90 |  | 36 |
| 38 | Provided sufficient time for the participants to reflect on their own experiences and listen to the experiences of other participants. | 4.60 | 1.00 | 1.00 |  | 37 |
| 39 | Responded to participants' comments with various techniques, such as providing frequent summaries (clarification), information (e.g., general ideas and examples of other participants), modest self-disclosure (the therapist's own experiences, thoughts, feelings, and behavior), or questions to broaden the participants' perspectives (e.g., gave extreme examples or asked about differences from past experiences). | 4.70 | 1.00 | 1.00 |  | 39 |
| **Focusing on key cognition or behaviors** |  |  |  |  |  |  |
| 40 | Conceptualized the cognition and behavior that were related to each participant's problems (e.g., depression, absence from work); had a working hypothesis. | 4.60 | 1.00 | 0.90 |  | 40 |
| 41 | Asked questions about cognition and behavior that were related to each participant's problems (e.g., depression, absence from work) to foster their understanding of themselves. | 4.50 | 1.00 | 1.00 |  | 41 |
| 42 | For participants who have difficulty in reflecting on their own cognitions, behaviours and feelings, support is tailored to their characteristics, for example, by providing them with options so that they can make their own choices. | 3.87 | 1.00 | 1.00 | Merged with 36 | 39 |
| 43 | Conceptualized participants' treatment tasks from their remarks and behavior in the session (e.g., attitudes toward the therapists and other participants, attitudes toward the programs, or homework adherence). | 3.87 | 1.00 | 1.00 |  | 42 |
| 44 | Explained to the participants the link between their personal treatment tasks and the program agenda. | 4.60 | 1.00 | 1.00 |  | 43 |
| 45 | When too much time was spent on the discussion on cognition and behavior of lower priority, refocused the attention on cognition and behavior of higher priority. | 4.90 | 1.00 | 1.00 |  | 44 |
| 46 | Identified a participant's cognitive-behavioral patterns that are common in different situations and identified underlying beliefs from background information such as their medical history, family status, and remarks at prior sessions | 4.70 | 0.60 | 0.90 | Split | 16<br>45 |
| **Strategy for change** |  |  |  |  |  |  |
| 47 | Depending on the characteristics of the participants and the objectives of the group, devise the way the programme is organised, including the choice of techniques, the order in which they are presented and the speed at which it progresses. | 4.70 | 0.80 | 0.90 | Merged with 48 | 46 |
| 48 | Chose techniques that were helpful to the participants based on their characteristics, such as their problems, motivation for the program, and comprehension abilities. | 4.80 | 1.00 | 1.00 |  | 46 |
| 49 | Explained how the chosen techniques can be helpful to each participant's treatment task. | 4.50 | 1.00 | 1.00 |  | 47 |

**Table 7.** Impact Score, CVR, CVI of G-CTS Items.

|  | Trial Items |  |  |  | Revision Method | Revised Items |
| --- | --- | --- | --- | --- | --- | --- |
| No | Therapist behavior | IS | CVR | CVI |  | No |
| **Application of cognitive-behavioral techniques** |  |  |  |  |  |  |
| 50 | Introduction (explanation): Explained the general usage of cognitive-behavioral techniques and their effectiveness. | 4.05 | 1.00 | 0.90 |  | 48 |
| 51 | Interventions: Applied cognitive-behavioral techniques to the problems of each participant and helped them to learn those techniques. | 3.87 | 1.00 | 1.00 |  | 52 |
| 52 | Summary: Discussed the results of using cognitive-behavioral techniques with the participants. | 3.78 | 1.00 | 1.00 |  | 49 |
| 53 | To enable participants to use the learned techniques outside of the session, the therapist will devise ways to apply the techniques, taking into account their practicability and motivation in their out-of-session life situations. | 4.40 | 1.00 | 0.90 | Split | 51<br>53 |
| 54 | Used examples and metaphors that were appropriate for the background and characteristics of the participants and adjust the adaptation of the technique. | 4.30 | 0.60 | 0.80 | Split | 50<br>46 |
| **Homework** |  |  |  |  |  |  |
| 55 | Assigned homework that is related to the agenda. | 4.80 | 1.00 | 0.90 |  | 54 |
| 56 | Devised homework so that the participants would not forget to do it. | 3.96 | 1.00 | 1.00 |  | 55 |
| 57 | Modified the homework to suit the understanding and situations of each participant. | 3.87 | 1.00 | 1.00 |  | 59 |
| 58 | Explained the rationale of the homework. ion. | 3.69 | 1.00 | 1.00 |  | 56 |
| 59 | Let the participants consider the feasibility of the homework and discussed possible barriers and ways to remove such barriers in advance. | 4.05 | 1.00 | 0.90 |  | 60 |
| 60 | Took some time to review the homework that was assigned in the last session. | 4.60 | 1.00 | 1.00 |  | 57 |
| 61 | Shared adherence and specific contents of the homework of each participant and their out-of-session life situations related homework among staff members in advance and utilized them for the progress of the session. | 4.50 | 0.80 | 0.80 | Split | 51<br>58 |
| 62 | Conceptualized the participants who have not done the homework. Discussed with such participants about possible modifications of the homework or ways to do it within their living conditions so that they can complete it. | 4.50 | 1.00 | 0.90 |  | 61 |
| **Intervention using relationships with other participants** |  |  |  |  |  |  |
| 63 | Fostered participants' behaviors that were helpful to other participants (e.g., empathy, finding positive points, speaking about common experiences, helping other participants solve problems). | 4.90 | 1.00 | 1.00 |  | 62 |
| 64 | Shared a particular participant's remarks with the whole group in a generalized form so that other participants could find the issue relevant to themselves. | 4.80 | 1.00 | 1.00 |  | 63 |
| 65 | Asked for responses (cognition, behavior, or emotions) of other participants in response to a specific participant's statement. Showed diversity. | 5.00 | 1.00 | 1.00 |  | 64 |
| 66 | Understood and responded to the participants' cognitions and emotions to others (e.g., when a participant felt, "I am not capable of doing the tasks that are easy for other participants," was sensitive to such participants and responded accordingly). | 4.80 | 1.00 | 1.00 |  | 65 |
| 67 | Made sure that each participant could participate in the program according to their level of awareness and ability (e.g., provided each participant with a role in which he/she could take advantage of their strength, or asked a question that the participant could respond with confidence). | 4.80 | 1.00 | 1.00 |  | 66 |

IS: Impact Score, CVR: Content Validity Ratio, CVI: Content Validity Index

### Study 2

The objective of this second study was to classify G-CBT therapists’ desirable behaviors according to the degree of difficulty in implementation of these conducts (beginner, intermediate, and advanced levels), thereby establishing a rating system for the Group Cognitive behavioral Therapy Scale (G-CTS).

## Methods

A survey was administered by the board members of the Japanese Association of Group Cognitive Behavioral Therapy. The clinical background of the respondents was diverse, consisting of fifteen clinical psychologists (42.9%), ten physicians specializing in psychiatry or psychosomatic medicine (28.8%), six psychiatric social workers (17.1%), three nurses (8.6%), and others (8.5%). Their areas of expertise were medical (n=26, 74.3%), welfare (n=6, 17.1%), education (n=3, 8.6%), industry (n=2, 5.7%), and judicial (n=2, 5.7%).

This survey asked the respondents to evaluate each point on the 66-item checklist by selecting one of the following five choices: 1) Beginner Level (Every G-CBT therapist is required to do this), 2) Intermediate Level (Skilled therapists are desired to do this), 3) Advanced Level (Skilled therapists may do this), 4) Too complex (It can be relevant with G-CBT, but is too difficult and even advanced-level therapists may not do this), and 5) Inappropriate (It is not relevant to G-CBT).

We classified each item into Beginner, Intermediate, and Advanced levels based on the response distribution and according to the following criteria:

1. The items with more than 10% of endorsement to “Advanced level” were categorized as “Advanced level”.
2. The items with <10% of endorsement to “Advanced level” and with more than 50% of endorsement to other specific difficulty level were categorized into that specific difficulty level.
3. The items with <10% of endorsement to “Advanced level” and with < 50% of endorsement to any other difficulty level were categorized as “Intermediate level”.

## Results

The results of the survey are shown in Tables 8 to 12. Twenty three items were classified as Beginner Level, thirty one as Intermediate Level, and twelve as Advanced Level. No item fell into “Too complex” or “Inappropriate”.

**Table 8.** Respondent Ratio of Checklist Items on Agenda Setting and Feedback.

|  |  | Response rate (%) |  |  |  | Difficulty level |
| --- | --- | --- | --- | --- | --- | --- |
| No | Therapist behavior | Beginner | Intermediate | Advanced | Inappropriate |  |
| Agenda setting |  |  |  |  |  |  |
| 1 | Clarified the agendas and the structure of the session to the participants (e.g., wrote the agendas and schedule on a whiteboard or printed material). | 82.9 | 14.3 | 2.9 | 0.0 | Beginner |
| 2 | Asked the participants about their impressions of and changes in their lives from the previous session, their current mood and physical conditions, and their implementation and impressions of their homework (check-in). | 74.3 | 22.9 | 2.9 | 0.0 | Beginner |
| 3 | Presented the agenda to the participants at the beginning of the session (usually within 5-10 minutes) and obtained their consent. | 74.3 | 20.0 | 0.0 | 5.7 | Beginner |
| 4 | Proceeded along with the set agenda. When changing the agenda, explained the rationale and obtain the participants' agreement. Intervened when a discussion deviates from the agenda or when the discussion pace was too slow. | 34.3 | 65.7 | 0.0 | 0.0 | Intermediate |
| 5 | Explained why the set agendas are useful/helpful to the participants. Modified the agenda as necessary, based on the feedback from the participants during the check-in and other related information (if any). | 14.3 | 65.7 | 20.0 | 0.0 | Advanced |
| 6 | Conceptualized the experiences of the participants and presented these to the participants by linking them to the agenda (e.g., when a participant said, "I can't get started easily although I know I have to do it," the therapist said, "Today's topic is 'behavioral activation,' a useful skill to deal with such problems"). | 22.9 | 60.0 | 17.1 | 0.0 | Advanced |
| Feedback |  |  |  |  |  |  |
| 7 | Checked participants' understanding and satisfaction <u>at the end of the session</u>. | 74.3 | 25.7 | 0.0 | 0.0 | Beginner |
| 8 | Received feedback from participants and shared with the participants what the therapist understood. | 74.3 | 25.7 | 0.0 | 0.0 | Beginner |
| 9 | Checked participants' understanding and satisfaction <u>throughout the session</u>. | 48.6 | 48.6 | 2.9 | 0.0 | Intermediate |
| 10 | Created an atmosphere that allowed the participants to express their moods and thoughts frankly, even if they were negative ones (e.g., asked a participant who remained silent during feedback and told them that even negative comments, if any, would be helpful for the group members to deepen their understanding; used self-disclosure techniques, such as asking, "The pacing of the session may have been a little too fast today. Did you find it difficult to follow?”). | 42.9 | 57.1 | 0.0 | 0.0 | Intermediate |
| 11 | Used the feedback from the participants to conceptualize the participants or modify the behavior of the therapist as needed. (e.g., 1: Slowed down when the session pace was perceived by the participants as being too fast. 2: In response to a specific participant’s comment, asked the whole group for comments in order to generalize that person’s comments, such as, “Thank you for your precious comment. It must have been a bit hard for you to share it. Are there any other people who have similar opinions?”) | 38.2 | 61.8 | 0.0 | 0.0 | Intermediate |
| 12 | Encouraged each participant to talk about their experiences in their own words (e.g., asked clarifying questions when a participant’s remark was too abstract). | 45.7 | 51.4 | 2.9 | 0.0 | Intermediate |

**Table 9.** Respondent Ratio of Checklist Items on Understanding, Interpersonal Effectiveness, and Collaboration.

|  |  | Response rate (%) |  |  |  | Difficulty level |
| --- | --- | --- | --- | --- | --- | --- |
| No | Therapist behavior | Beginner | Intermediate | Advanced | Inappropriate |  |
| Understanding |  |  |  |  |  |  |
| 13 | Listened to each participant and understood their thoughts and emotions accurately. | 65.7 | 28.6 | 5.7 | 0.0 | Beginner |
| 14 | Elicited remarks from each participant, understood their cognition, behavior, and emotions, and conveyed the therapist's understanding to the participants (case conceptualization). | 25.7 | 65.7 | 5.7 | 2.9 | Intermediate |
| 15 | Tried to understand the participants' moods through verbal and non-verbal communication and to convey the therapist's understandings through his/her attitudes and behaviors (understanding). | 42.9 | 48.6 | 8.6 | 0.0 | Intermediate |
| 16 | Understood the dynamics and processes taking place in the group and shared them with the co-leader(s) (e.g., paid attention to the occurrence of subgroups and fixed the roles of the participants). | 34.3 | 57.1 | 8.6 | 0.0 | Intermediate |
| 17 | Understood the participants' thoughts and feelings, not only from the information in the group session, but also from background information such as their medical history, family status, and remarks at prior sessions, and shared these with the participants (case conceptualization). | 11.4 | 65.7 | 20.0 | 2.9 | Advanced |
| Interpersonal effectiveness |  |  |  |  |  |  |
| 18 | Was fair and honest with all participants. Didn't take condescending or deliberately humble attitudes. Did not evade participants' questions. Conveyed warmth and interest to each participant not only by the contents of his/her remarks but also with nonverbal behavior such as tone of voice and eye contact. | 68.6 | 28.6 | 2.9 | 0.0 | Beginner |
| 19 | Appropriately intervened with participants' behavior that might have hindered other participants' sense of security (e.g., competition, aggression, and imposition of opinions on others). | 54.3 | 42.9 | 2.9 | 0.0 | Beginner |
| 20 | Made the group a safe and secure place by balancing attending to each participant with managing the group as a whole. | 62.9 | 34.3 | 2.9 | 0.0 | Beginner |
| 21 | Showed confidence that the therapist was capable of helping the participants (e.g., showed expertise related to the program or illustrated examples of thoughts and feelings that are common to many of the participants). Was not distant towards the participants. Did not show cold or intimidating attitudes. | 31.4 | 62.9 | 5.7 | 0.0 | Intermediate |
| 22 | Was aware of the therapist's own emotions during the session and verbalized or used them to understand the participants' feelings. Provided appropriate self-disclosure as necessary. | 14.3 | 68.6 | 14.3 | 2.9 | Advanced |
| **Collaboration** |  |  |  |  |  |  |
| 23 | In response to the remarks by one participant, asked other participants or the co-leader whether they had similar experiences and thereby elicited empathy and opinions. | 57.1 | 37.1 | 5.7 | 0.0 | Beginner |
| 24 | Informed the participants that their active participation was essential for the progress of the therapy (e.g., addressed a participant's reluctance to take part in the tasks in the session or homework ). | 48.6 | 48.6 | 2.9 | 0.0 | Intermediate |
| 25 | Considered sharing the contents of a participant's remarks with the others so that the input could be treated as common issues in the entire group. Did not end with a 1:1 conversation between therapist and participant. | 48.6 | 45.7 | 5.7 | 0.0 | Intermediate |
| 26 | Facilitated the session by cooperatively sharing roles among the therapists – i.e., moderator, follower, confrontational. | 45.7 | 45.7 | 5.7 | 2.9 | Intermediate |
| 27 | Explained the rationale for how the program can be helpful for each participant's treatment goals. | 42.9 | 45.7 | 11.4 | 0.0 | Advanced |

**Table 10.** Respondent Ratio of Checklist Items on Pacing and Efficient Use of Time and Guided Discovery.

|  |  | Response rate (%) |  |  |  | Difficulty level |
| --- | --- | --- | --- | --- | --- | --- |
| No | Therapist behavior | Beginner | Intermediate | Advanced | Inappropriate |  |
| **Pacing and efficient use of time** |  |  |  |  |  |  |
| 28 | Informed the participants that the time of the sessions should be shared equally among the participants. Clarified as a rule that one participant should not speak too long. | 77.1 | 20.0 | 2.9 | 0.0 | Beginner |
| 29 | Presented a rough time allocation for each agenda. | 74.3 | 22.9 | 2.9 | 0.0 | Beginner |
| 30 | Progressed the session according to the pre-planned schedule. | 28.6 | 68.6 | 2.9 | 0.0 | Intermediate |
| 31 | Focused on important topics. When the group was distracted from the central topics and the discussion became unproductive, gently interrupted the discussion and led the discussion back to the original topic. | 42.9 | 54.3 | 2.9 | 0.0 | Intermediate |
| 32 | Made the discussion fruitful. When important treatment issues were not sufficiently discussed (e.g., a participant remained silent or was dedicated to unproductive chatting), the therapist cast questions to facilitate deeper discussion or explored the reason the participant(s) was not facing his/her own problems. | 22.9 | 60.0 | 17.1 | 0.0 | Intermediate |
| 33 | Encouraged participants to use their time equally. When a participant spent too much time on him/herself, conceptualized the participant and limited his/her talk (e.g., gently interrupted him/her by providing a summary). | 17.1 | 71.4 | 11.4 | 0.0 | Intermediate |
| 34 | Adjusted the session pace according to the characteristics of the participants. For hurried participants, instructed them to pace down and listen more carefully to other participants. Conceptualized and attended to the participants who were not motivated enough or who had difficulty in understanding the program. | 5.7 | 80.0 | 14.3 | 0.0 | Intermediate |
| **Guided discovery** |  |  |  |  |  |  |
| 35 | Asked specific questions to help participants identify their cognition and behavior (e.g., when asking patients to identify automatic thoughts, directed them to think about specific situations using 5W1H). | 71.4 | 28.6 | 0.0 | 0.0 | Beginner |
| 36 | Did not confront the participants. Avoided pressing or confronting issues that they were not fully aware of or were denying. | 65.7 | 34.3 | 0.0 | 0.0 | Beginner |
| 37 | Provided sufficient time for the participants to reflect on their own experiences and listen to the experiences of other participants. | 60.0 | 37.1 | 2.9 | 0.0 | Beginner |
| 38 | Provided support that was tailored to each participant depending on their characteristics (e.g., for those who found it difficult to recognize their cognition, behavior, or emotions, the therapist offered clues, such as providing choices that the participants may pick up). | 20.0 | 68.6 | 11.4 | 0.0 | Intermediate |
| 39 | Responded to participants' comments with various techniques, such as providing frequent summaries (clarification), information (e.g., general ideas and examples of other participants), modest self-disclosure (the therapist's own experiences, thoughts, feelings, and behavior), or questions to broaden the participants' perspectives (e.g., gave extreme examples or asked about differences from past experiences). | 2.9 | 82.9 | 14.3 | 0.0 | Intermediate |

**Table 11.**
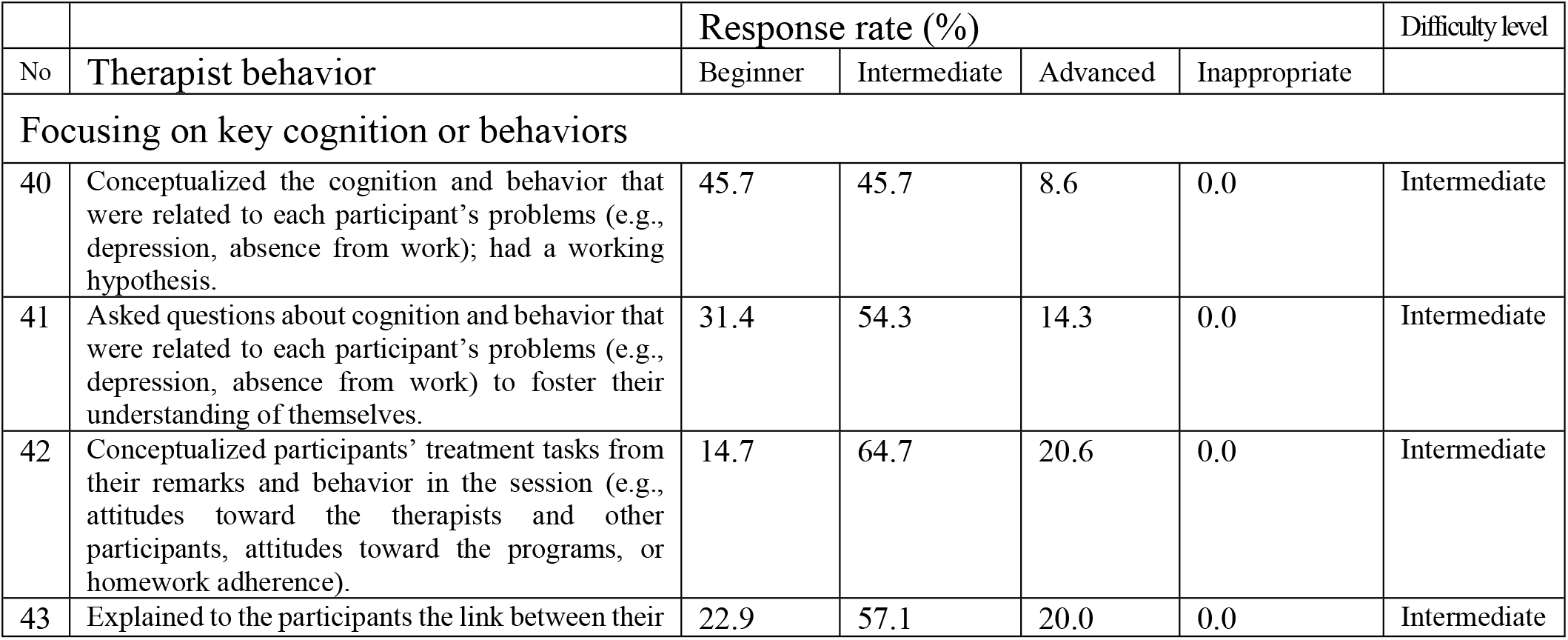
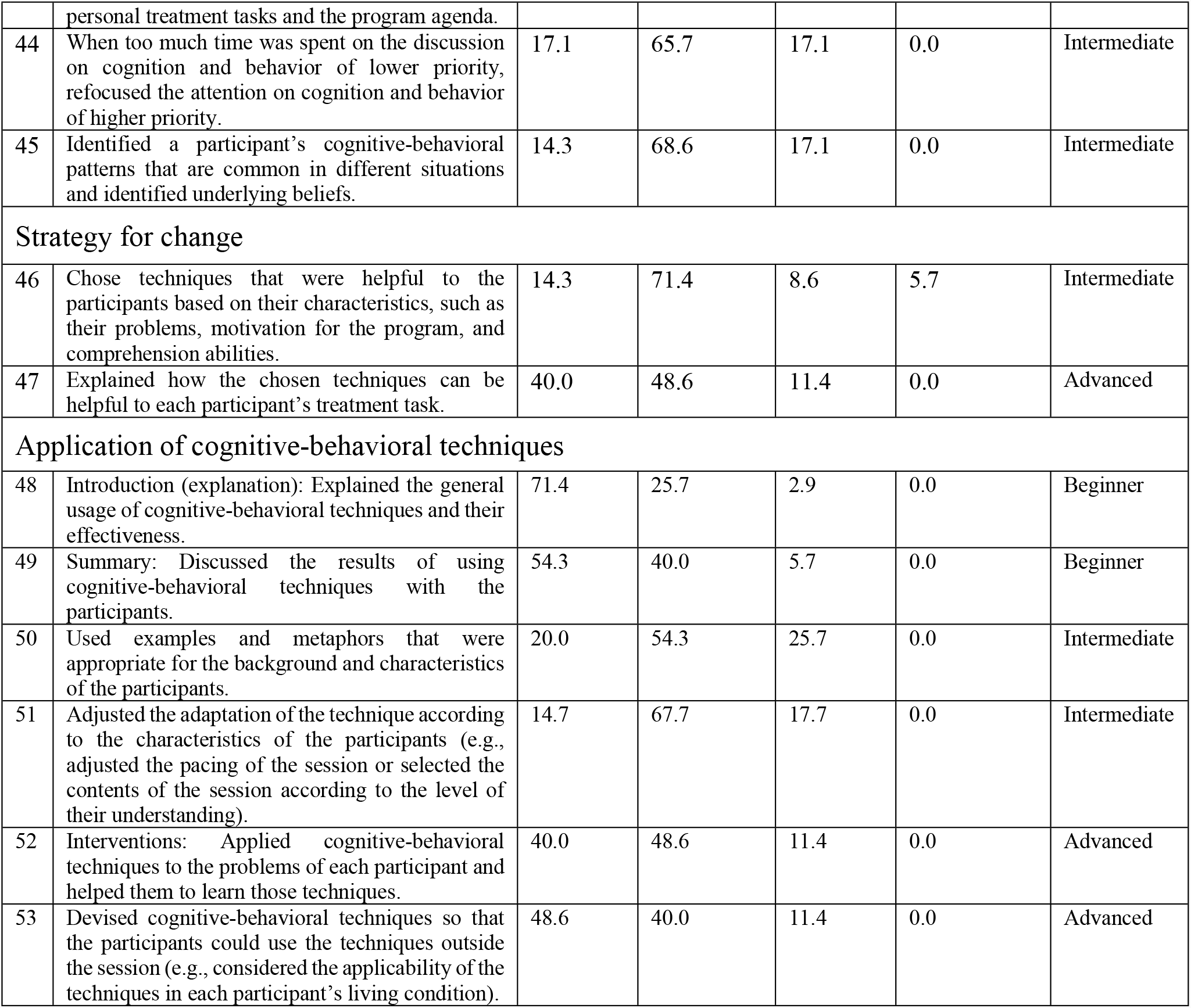
Respondent Ratio of Checklist Items on Focusing on Key Cognition or Behaviors, Strategy for Change, and Application of Cognitive-Behavioral Techniques.

|  |  | Response rate (%) |  |  |  | Difficulty level |
| --- | --- | --- | --- | --- | --- | --- |
| No | Therapist behavior | Beginner | Intermediate | Advanced | Inappropriate |  |
| Focusing on key cognition or behaviors |  |  |  |  |  |  |
| 40 | Conceptualized the cognition and behavior that were related to each participant's problems (e.g., depression, absence from work); had a working hypothesis. | 45.7 | 45.7 | 8.6 | 0.0 | Intermediate |
| 41 | Asked questions about cognition and behavior that were related to each participant's problems (e.g., depression, absence from work) to foster their understanding of themselves. | 31.4 | 54.3 | 14.3 | 0.0 | Intermediate |
| 42 | Conceptualized participants' treatment tasks from their remarks and behavior in the session (e.g., attitudes toward the therapists and other participants, attitudes toward the programs, or homework adherence). | 14.7 | 64.7 | 20.6 | 0.0 | Intermediate |
| 43 | Explained to the participants the link between their personal treatment tasks and the program agenda. | 22.9 | 57.1 | 20.0 | 0.0 | Intermediate |
| 44 | When too much time was spent on the discussion on cognition and behavior of lower priority, refocused the attention on cognition and behavior of higher priority. | 17.1 | 65.7 | 17.1 | 0.0 | Intermediate |
| 45 | Identified a participant's cognitive-behavioral patterns that are common in different situations and identified underlying beliefs. | 14.3 | 68.6 | 17.1 | 0.0 | Intermediate |
| **Strategy for change** |  |  |  |  |  |  |
| 46 | Chose techniques that were helpful to the participants based on their characteristics, such as their problems, motivation for the program, and comprehension abilities. | 14.3 | 71.4 | 8.6 | 5.7 | Intermediate |
| 47 | Explained how the chosen techniques can be helpful to each participant's treatment task. | 40.0 | 48.6 | 11.4 | 0.0 | Advanced |
| **Application of cognitive-behavioral techniques** |  |  |  |  |  |  |
| 48 | Introduction (explanation): Explained the general usage of cognitive-behavioral techniques and their effectiveness. | 71.4 | 25.7 | 2.9 | 0.0 | Beginner |
| 49 | Summary: Discussed the results of using cognitive-behavioral techniques with the participants. | 54.3 | 40.0 | 5.7 | 0.0 | Beginner |
| 50 | Used examples and metaphors that were appropriate for the background and characteristics of the participants. | 20.0 | 54.3 | 25.7 | 0.0 | Intermediate |
| 51 | Adjusted the adaptation of the technique according to the characteristics of the participants (e.g., adjusted the pacing of the session or selected the contents of the session according to the level of their understanding). | 14.7 | 67.7 | 17.7 | 0.0 | Intermediate |
| 52 | Interventions: Applied cognitive-behavioral techniques to the problems of each participant and helped them to learn those techniques. | 40.0 | 48.6 | 11.4 | 0.0 | Advanced |
| 53 | Devised cognitive-behavioral techniques so that the participants could use the techniques outside the session (e.g., considered the applicability of the techniques in each participant's living condition). | 48.6 | 40.0 | 11.4 | 0.0 | Advanced |

**Table 12.** Respondent Ratio of Checklist Items on Homework and Intervention Using Relationships with Other Participants.

|  |  | Response rate (%) |  |  |  | Difficulty level |
| --- | --- | --- | --- | --- | --- | --- |
| No | Therapist behavior | Beginner | Intermediate | Advanced | Inappropriate |  |
| Homework |  |  |  |  |  |  |
| 54 | Assigned homework that is related to the agenda. | 71.4 | 25.7 | 2.9 | 0.0 | Beginner |
| 55 | Devised homework so that the participants would not forget to do it. | 74.3 | 22.9 | 2.9 | 0.0 | Beginner |
| 56 | Explained the rationale of the homework. | 68.6 | 25.7 | 5.7 | 0.0 | Beginner |
| 57 | Took some time to review the homework that was assigned in the last session. | 85.7 | 11.4 | 2.9 | 0.0 | Beginner |
| 58 | Shared adherence and specific contents of the homework of each participant among staff members in advance and utilized them for the progress of the session. | 74.3 | 20.0 | 5.7 | 0.0 | Beginner |
| 59 | Modified the homework to suit the understanding and situations of each participant. | 45.7 | 45.7 | 8.6 | 0.0 | Intermediate |
| 60 | Let the participants consider the feasibility of the homework and discussed possible barriers and ways to remove such barriers in advance. | 45.7 | 40.0 | 14.3 | 0.0 | Advanced |
| 61 | Conceptualized the participants who have not done the homework. Discussed with such participants about possible modifications of the homework or ways to do it within their living conditions so that they can complete it. | 45.7 | 40.0 | 11.4 | 2.9 | Advanced |
| Intervention using relationships with other participants |  |  |  |  |  |  |
| 62 | Fostered participants' behaviors that were helpful to other participants (e.g., empathy, finding positive points, speaking about common experiences, helping other participants solve problems). | 62.9 | 34.3 | 2.9 | 0.0 | Beginner |
| 63 | Shared a particular participant's remarks with the whole group in a generalized form so that other participants could find the issue relevant to themselves. | 48.6 | 42.9 | 8.6 | 0.0 | Intermediate |
| 64 | Asked for responses (cognition, behavior, or emotions) of other participants in response to a specific participant's statement. Showed diversity. | 45.7 | 51.4 | 2.9 | 0.0 | Intermediate |
| 65 | Understood and responded to the participants' cognitions and emotions to others (e.g., when a participant felt, "I am not capable of doing the tasks that are easy for other participants," was sensitive to such participants and responded accordingly). | 20.0 | 68.6 | 11.4 | 0.0 | Advanced |
| 66 | Made sure that each participant could participate in the program according to their level of awareness and ability (e.g., provided each participant with a role in which he/she could take advantage of their strength, or asked a question that the participant could respond with confidence). | 25.7 | 62.9 | 11.4 | 0.0 | Advanced |

Additionally, we reorganized the above checklist for group therapists’ desirable behaviors according to the Revised Cognitive Therapy Scale (CTS-R) rating system (18), which provides anchor points for evaluation on the seven-level Likert scale. We did so because both the level of competence in the execution of CBT techniques and adherence to CBT protocols are taken into consideration in the CTS-R. A score of 0 indicates non-adherence to a standard CBT protocol, and a score of 6 indicates extreme expertise even in difficult cases.

We added the following scoring rules based on the behavioral checklists:

1. Three points or more are given if a therapist satisfied all the Beginner-level items.
2. Four points or more are given if a therapist satisfies all the Beginner-level and Intermediate-level items.
3. Five points or more are given if a therapist satisfies all the items of both the Beginner-level and Intermediate-level items, in addition to some of the Advanced-level items (Table 13).

**Table 13.**
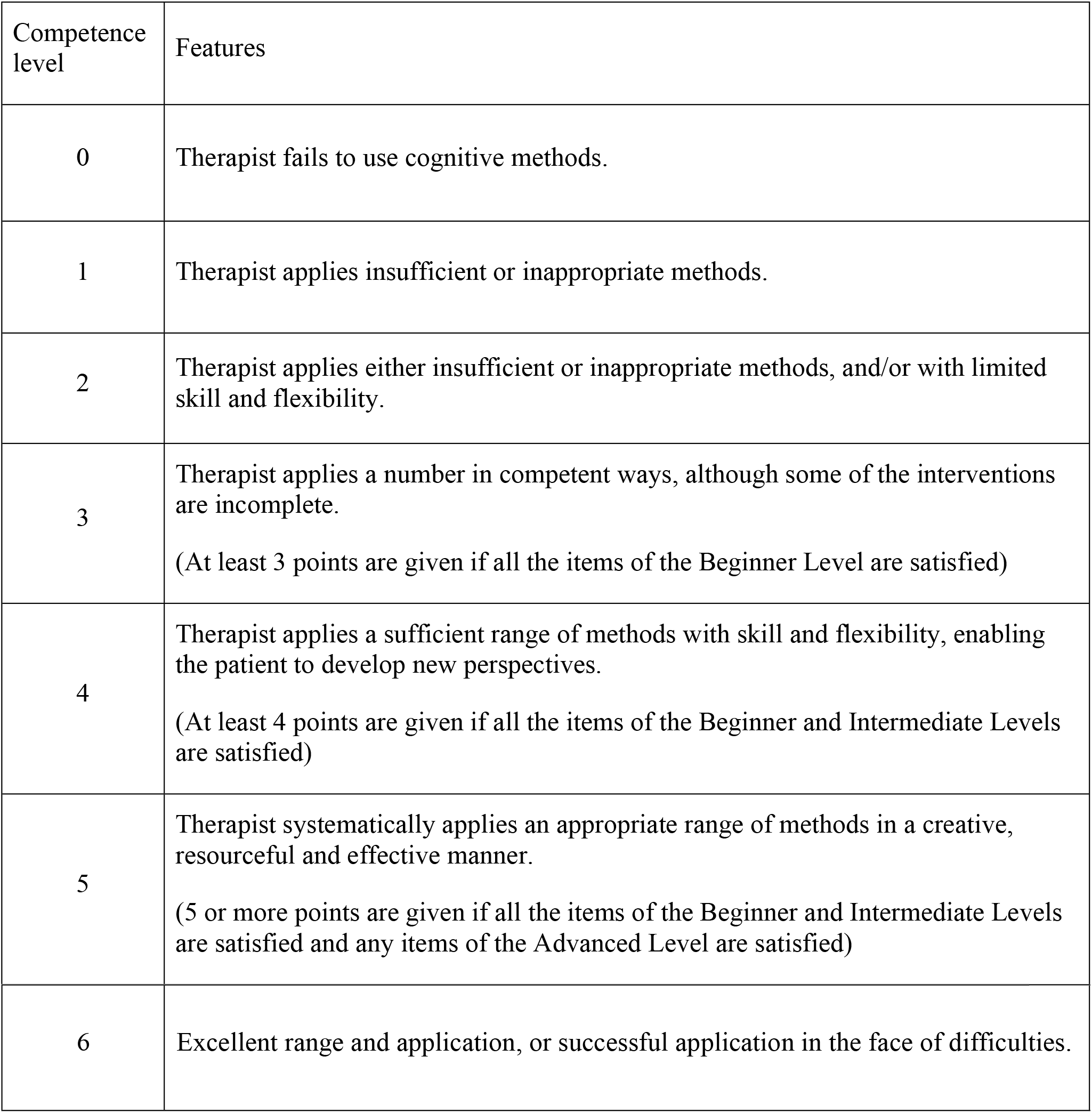
Scoring for each item.

| Competence level | Features |
| --- | --- |
| 0 | Therapist fails to use cognitive methods. |
| 1 | Therapist applies insufficient or inappropriate methods. |
| 2 | Therapist applies either insufficient or inappropriate methods, and/or with limited skill and flexibility. |
| 3 | Therapist applies a number in competent ways, although some of the interventions are incomplete.<br>(At least 3 points are given if all the items of the Beginner Level are satisfied) |
| 4 | Therapist applies a sufficient range of methods with skill and flexibility, enabling the patient to develop new perspectives.<br>(At least 4 points are given if all the items of the Beginner and Intermediate Levels are satisfied) |
| 5 | Therapist systematically applies an appropriate range of methods in a creative, resourceful and effective manner.<br>(5 or more points are given if all the items of the Beginner and Intermediate Levels are satisfied and any items of the Advanced Level are satisfied) |
| 6 | Excellent range and application, or successful application in the face of difficulties. |

### Study 3

The objective of this third study was to investigate the reliability and validity of the Group Cognitive Therapy Scale (G-CTS). We used two groups of video samples - training sessions conducted by novice therapists (Beginner Group samples) and by skilled therapists (Advanced Group samples).

A past study that examined degree of proficiency of group CBT therapists’ skills yielded moderate effect size (Cohen’s *d* = 0.58) (18). Since our study sample comprised expert- and novice-therapists, we assumed that the effect size will be large (*d* = 0.8). Using the G*Power 3 program (28), we estimated our target sample size as 23 participants per arm to detect an effect size of 0.8 at an alpha error level of 0.05 and a beta error level of 0.2.

## Methods

### Samples

Two sets of G-CBT session videos were prepared (Beginner Group samples and Advanced Group samples). The Beginner Group videos were video-recordings of the roleplays of typical G-CBT sessions for mild depression, conducted by psychology graduate students as a way of training. The Advanced Group videos were video-recordings of actual clinical G-CBT sessions for patients with mild depression in actual clinical therapy, as the Beginner Group role-played with typical minor depression patients. The therapists in Advanced Group completed the basic G-CBT training course offered by the Association of Cognitive Behavioral Group Therapy in Japan (https://cbgt.org/) and had five or more years of experience in G-CBT (mean length of G-CBT experience = 10.47 years). We aimed to prepare 23 videos for each group, but for a practical reason, we were able to shoot only 18 videos for the Beginner Group.

### Participants

Two clinicians with five or more years of experience in G-CBT who had undergone training in G-CTS (a two-hour lecture and an evaluation exercise) independently rated each video, based on the G-CTS.

### Statistical analysis

We computed Cronbach’s alpha coefficient to examine the internal consistency of the G-CTS and interclass correlation coefficients to assess the inter-rater reliability. Since the data did not show normal distribution, we compared the total G-CTS scores of the Beginner Group and the Advanced Group sessions using the Mann-Whitney test, to examine predictive validity.

## Results

### Internal consistency

Cronbach’s alpha coefficients were 0.95[.93−.97] and 0.96[.95−.98] for each rater, confirming a high level of internal consistency.

### Inter-item correlations

We computed correlation coefficients between each item, which ranged between .55 and .83, showing that the redundancy was not very high.

### Inter-rater reliability

Interclass correlation coefficients for each item were satisfactory, ranging from 0.75 to 0.90. The interclass correlation coefficient for the total G-CTS score was 0.82 [.68−.90] (Table 14).

**Table 14.** Interclass Correlation Coefficients for Pairs of Raters for Each Item in the G-CTS.

| Item |  | Interclass<br>Correlation<br>Coefficients | Confidence Interval<br>(95%) |
| --- | --- | --- | --- |
| 1 | Agenda setting | .82 | .68-.90 |
| 2 | Feedback | .81 | .66-.89 |
| 3 | Understanding | .81 | .67-.90 |
| 4 | Interpersonal effectiveness | .80 | .66-.89 |
| 5 | Collaboration | .81 | .66-.89 |
| 6 | Pacing and efficient use of time | .75 | .58-.86 |
| 7 | Guided discovery | .77 | .60-.87 |
| 8 | Focusing on key cognition or behaviors | .78 | .63-.88 |
| 9 | Strategy of change | .77 | .61-.87 |
| 10 | Application of cognitive-behavioral techniques | .90 | .82-.95 |
| 11 | Homework | .88 | .79-.94 |
| 12 | Intervention using relationships with other participants | .80 | .65-.89 |
|  | Total | .82 | .68-.90 |

### Predictive validity

The comparison between each G-CTS score of the Beginner Group and Advanced Group videos is shown in Table 15. All the scores of each item and the total were significantly higher in the Advanced Group than in the Beginner Group, with large effect size [.60−.88].

**Table 15.**
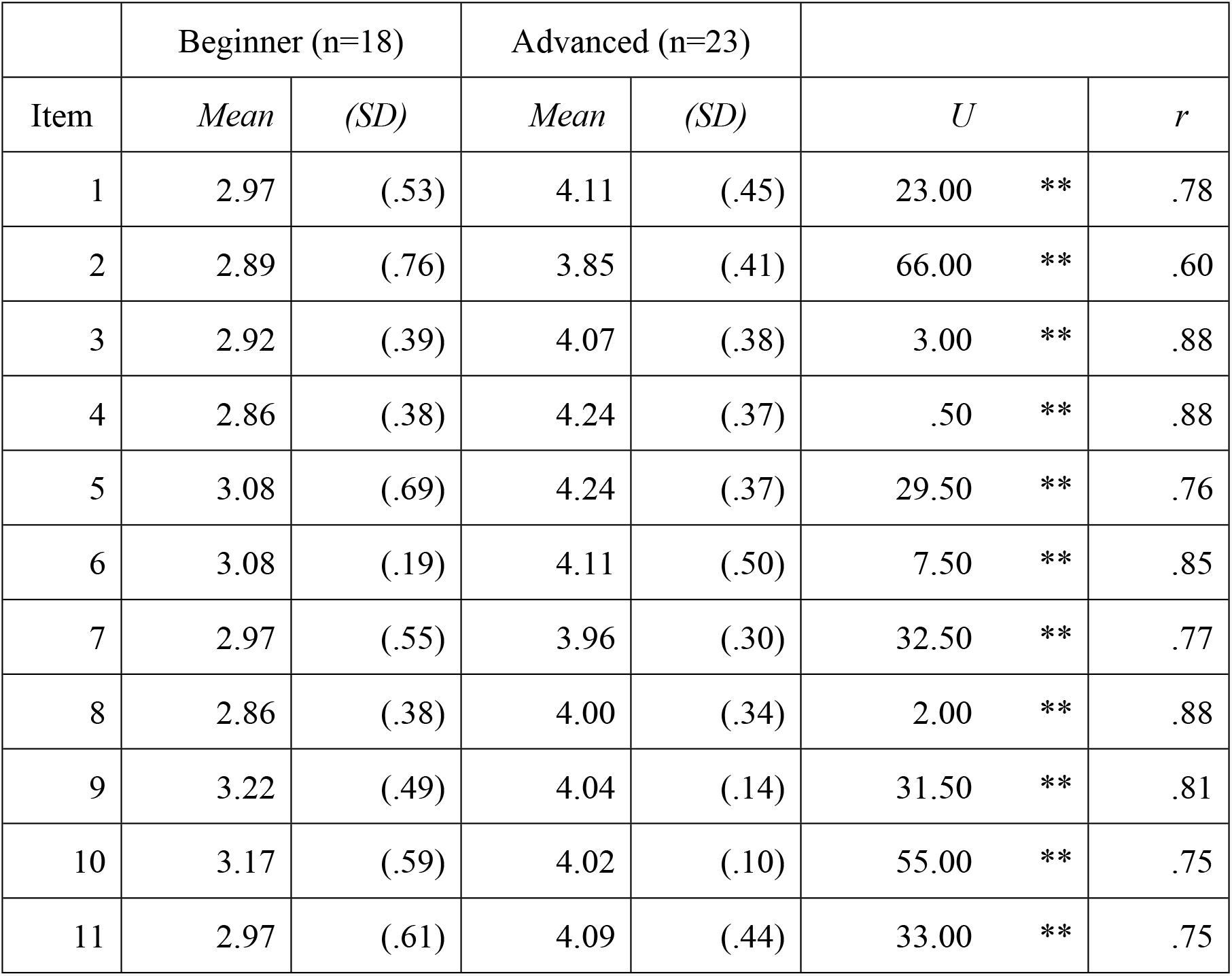
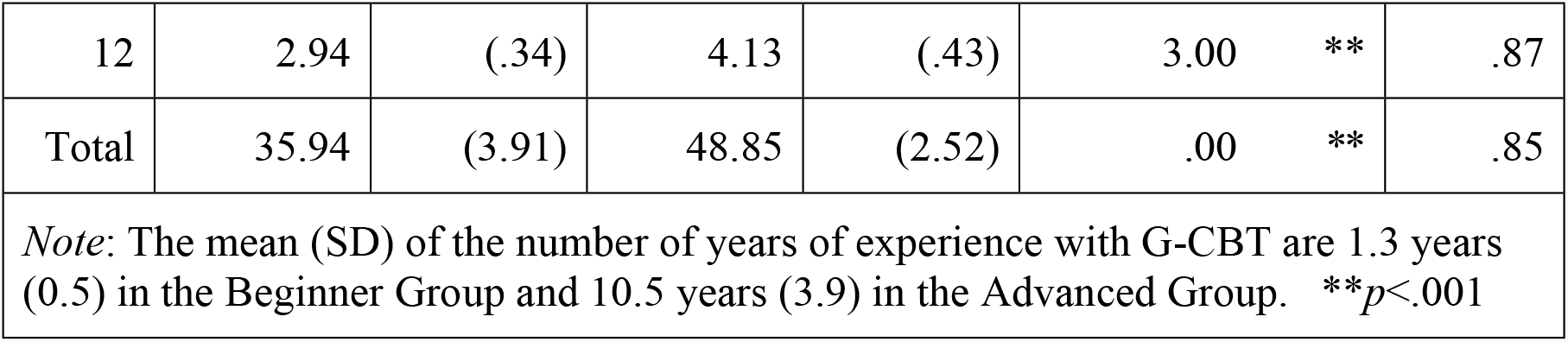
Differences in G-CTS Scores According to Number of Years of Experience.

## Discussion

The purpose of this study was to develop a scale for evaluating the quality of G-CBT and to investigate its validity and reliability.

We created a twelve-item scale for G-CBT with solid behavioral anchors by reforming the CTS, which is a well-established scale to evaluate the quality of individual CBT, and by adding specific skills for group psychotherapy. The most important feature of the G-CTS was that it increased rater agreement by describing many specific examples of desired therapist behavior.

We demonstrated that G-CTS has high internal reliability and high inter-rater reliability compared with existing instruments that measure the quality of individual CBT. For example, the interclass correlation coefficients of the CTS-R and Assessment of Core CBT Skills (ACCS) were 0.40-0.86 and 0.27-0.83, respectively (18, 29). The interclass correlation coefficients for the total scores of CTS-Rs were 0.63 (thirteen-item edition) and 0.57 (fourteen-item edition) (18). We conclude that our scale achieved an exceptionally high inter-rater reliability because it has clear behavioral anchors for rating. Further, the high predictive validity of the G-CTS was demonstrated by comparing the total scores and the scores for each item in the Beginner Group and the Advanced Group of G-CBT therapists.

The G-CTS behavioral checklist created in this study provides concrete guidelines that can be used by therapists to hone their skills in G-CBT. The G-CTS checklist and the rating scale offer a framework that can be utilized in teaching, supervision, research, and qualification of G-CBT therapists as well as in programs that facilitate education and training of new practitioners and contribute to the dissemination of G-CBT. Classifying therapist behaviors in this manner not only increases the convenience of administering the scale but also clearly indicates priorities for skill acquisition for beginner practitioners and supervisors of G-CBT.

There are several limitations to this study. First, the video sessions used in the assessment were exclusively of mild-depression patients, and the reliability and validity of the G-CTS was not examined for G-CBT in other disorders. Second, G-CTS targets the only leader-therapist and does not include co-leaders. Third, there were substantial differences in the videos of Beginner and Advanced Groups - role plays were filmed in the beginner videos, and clinical scenes were filmed in advanced videos – so it may be inappropriate to compare the two. Fourth, since the therapists’ years of experience and ages were relatively proportional, the video evaluators may have been able to predict the years of experience from the therapists’ physical appearances. Lastly and most importantly, the validation process was limited to internal consistency, inter-rater reliability and predictive validity. We were not able to conduct factor analysis due to small sample size. Further verification with larger sample is needed.

Despite these limitations, our study is noteworthy since we developed a novel rating scale for G-CBT that is not specific to a certain disorder. The scale has solid behavioral anchors, which led to high reliability of the scale.

Future research implications include the following: First, the reliability and validity of the G-CTS may need to be verified in samples other than depression. It is also necessary to establish an evaluation system that can accommodate the evaluation of co-therapists. Finally, since therapists’ desirable behavior may be different among different cultures, cultural adaptation of behavioral anchors of the G-CTS may be needed.

## Data Availability

Data are available from the authors for researchers who meet the criteria for access to confidential data.

## Author Contributions

M.N., M.M., M.O., H.K., and D.F. designed the study. M.N., M.M., and D.F. wrote the manuscript. Data analyses were performed by M.N. under supervision of H.K., D.F., and M.O. provided clinical advice. M.N. and M.M. provided their sessions’ video data. M.N. and M.M. reviewed the literature. M.N., M.M., and M.O. rated the videos. All authors contributed to the article and approved the submitted version.

## Acknowledgments

We thank Tsuyoshi Akiyama, and study group of group cognitive behavior therapy for their advice on scale creation and for their cooperation to our research.

## Ethics Approval and informed consent

This study was reviewed and approved by the Ethics Committee of the National Hospital Organization Hizen Psychiatric Medical Center (approval number: 28-7). The participants provided their written informed consent to participate in this study.

## References

1. Patterson F, Fleming J and Doig E. Group-based delivery of interventions in traumatic brain injury rehabilitation: a scoping review. Disabil Rehabil (2016) 38: 1961–1986. doi: 10.3109/09638288.2015.1111436

2. Tucker M, Oei T. Is group more cost effective than individual cognitive behaviour therapy? The evidence is not solid yet. Behav Cogn Psychother (2007) 35: 77–91. doi: 10.1017/S1352465806003134

3. National Institute for Health and Clinical Excellence: NICE Depression: Evidence Update April 2012: A Summary of Selected New Evidence Relevant to NICE Clinical Guideline 90 ‘The Treatment and Management of Depression in Adults.’ (2009)

4. Yoon I, Slade K, Fazel S. Outcomes of psychological therapies for prisoners with mental health problems: A systematic review and meta-analysis. J Consult Clin Psychol (2017) 85: 783–802. doi: 10.1037/ccp0000214

5. Mahon L, Leszcz M. The Interpersonal Model of Group Psychotherapy. Int J Group Psychoth (2017) 67: S121–S130. doi: org/10.1080/00207284.2016.1218286

6. Bieling PJ, Mccabe RE, Antony MM. Cognitive-behavioral Therapy in Groups (2006) The Guilford Press. doi: 10.1002/acp.1384

7. Barlow SH. A strategic three-year plan to teach beginning, intermediate, and advanced group skills. J Spec Group Work (2010) 29: 113–126.

8. Folmo EJ, Karterud SW, Bremer K, Walther KL, Kvarstein EH, Pedersen GA. The design of the MBT-G adherence and quality scale. Scand J Psychol (2017) 58: 341–349. doi: 10.1111/sjop.12375.

9. Karterud S. Mentalization-based Group Therapy (MBT-G): A Theoretical, Clinical and Research Manual. (2015) London: Oxford University Press.

10. Decker SD, Nich C, Carroll KM, Martino S. Development of the Therapist Empathy Scale. Behav Cogn Psychother (2014) 42: 339–354. doi:10.1017/S1352465813000039.

11. Dobson KS, & Singer AR. Definitional and practical issues in the assessment of treatment integrity. Clin Psychol (New York) (2005) 12: 384–387. doi: 10.1093/clipsy.bpi046

12. Laireiter AR, Willutzki U. Self-reflection and self-practice in training of cognitive behaviour therapy: An overview. Clin Psychol Psychother (2003) 10: 19–30. doi: 10.1002/cpp.348

13. McHugh RK, Barlow DH. The dissemination and implementation of evidence-based psychological treatments: A review of current efforts. Am Psychol (2010) 65: 73–84. doi: 10.1037/a0018121

14. Weck F, Bohn C, Ginzburg DM, Ulrich S. Treatment integrity: Implementation, assessment, evaluation and correlations with outcome. Verhaltenstherapie (2011) 21: 99–107. doi:10.1159/000328840.

15. Hepner KA, Stern S, Paddock SM, Hunter SB, Osilla KC, Watkins KE. A fidelity coding guide for a group cognitive behavioral therapy for depression. RAND Corporation. (2011) http://www.rand.org/pubs/technical_reports/TR980.html

16. Wong D, Grace N, Baker K, McMahon G. Measuring clinical competencies in facilitating group-based rehabilitation interventions: development of a new competency checklist. Clin Rehabil (2019) 33: 1079–1087. doi: 10.1177/0269215519831048.

17. Young JE, Beck AT. Cognitive Therapy Scale: Rating Manual. (1980) Unpublished Manuscript, University of Pennsylvania.

18. Blackburn I, James IA, Milne DL, Baker C, Standart S, Garland A, et al. The revised cognitive therapy scale (CTS-R): psychometric properties. Behav Cogn Psychoth (2001) 29: 431–446. doi: 10.1017/S1352465801004040

19. Elkin I, Parloff MB, Hadley SW, Autry, JH. NIMH Treatment of Depression Collaborative Research Program. Background and research plan. Arch gen psychiatry (1985) 42: 305–316. doi: 10.1001/archpsyc.1985.01790260103013

20. James IA, Blackburn IM, Milne DL, Reichfelt FK. Moderators of trainee therapists’ competence in cognitive therapy. Br J Health Psychol (2001) 40: 131–141. doi: 10.1348/014466501163580

21. Polit DF, Beck CT. Resource manual for nursing research: generating and assessing evidence for nursing practice. Wolters Kluwer Health: Lippincott Williams &Wilkins (2012).

22. Juniper EF, Guyatt GH, Streiner DL, King DR. Clinical impact versus factor analysis for quality of life questionnaire construction. J Clin Epidemiol. (1997) 50: 233–238. doi: 10.1016/S0895-4356(96)00377-0.

23. Lawshe CH. A quantitative approach to content validity. Pers Psychol. (1975) 28:563–575.

24. American Group Psychotherapy Association. Clinical Practice Guideline for Group Psychotherapy (2007) https://www.agpa.org/home/practice-resources/practice-guidelines-for-group-psychotherapy

25. White JR. Depression. In White JR, Freeman AS. (Eds.), Cognitive Behavioral Group Therapy: For Specific Problems and Populations (2000) American Psychological Association.

26. Wagner CC, Ingersoll KS. Motivational Interviewing in Groups (2013) New York: The Guilford Press.

27. Lieberman MA, Miles MB, Yalom ID. Encounter Groups: First Facts. (1973) New York: BasicBooks.

28. Faul F, Erdfelder E, Lang A G, & Buchner A. G*Power 3: A flexible statistical power analysis program for the social, behavioral, and biomedical sciences. Behavior Research Methods (2007) 39, 175–191. https://doi.org/10.3758/BF03193146

29. Muse K, McManus F, Rakovshik S, Thwaites R. Development and psychometric evaluation of the assessment of core CBT skills (ACCS): An observation-based tool for assessing cognitive behavioral therapy competence. Psychol Assess (2017) 29: 542–544. doi: 10.1037/pas0000372

30. Burlingame GM, Barlow SH. Outcome and process differences between professional and nonprofessional therapists in time-limited group psychotherapy. Int J Group Psychother (1996) 46: 455–478. doi: org/10.1080/00207284.1996.11491505

31. Schnur JB, Montgomery GH. A systematic review of therapeutic alliance, group cohesion, empathy, and goal consensus/collaboration in psychotherapeutic interventions in cancer: Uncommon factors? Clin Psychol Rev (2010) 30: 238–247. doi: 10.1016/j.cpr.2009.11.005.

32. Rice A. Common therapeutic factors in bereavement groups. Death Stud (2015) 39: 165–172. doi: 10.1080/07481187.2014.946627.

33. Gallagher ME, Tasca GA, Ritchie K, Balfour L, Maxwell H. Interpersonal learning is associated with improved self-esteem in group psychotherapy for women with binge eating disorder. Psychotherapy (2014) 51: 66–77. doi: org/10.1037/a0031098

34. Ahmed SA, Mona R, Salwa E, Rania M. Therapeutic Factors in Group Psychotherapy: A Study of Egyptian Drug Addicts. J Groups Addict Recover (2010) 5: 194–213. doi: org/10.1080/1556035X.2010.523345

35. Kennard D. Review of Therapeutic Factors in Group Psychotherapy. Group Analysis (1987) 20: 179–180. doi: org/10.1177/0533316487202013

33. Bloch S, Crouch E, Reibstein J. Therapeutic factors in group psychotherapy. A review. Arch gen psychiatry (1981) 38: 519–526. doi: 10.1001/archpsyc.1980.01780300031003.

34. Barlow SH, Burlingame GM, Fuhriman A. Therapeutic applications of groups: From Pratt’s “thought control classes” to modern group psychotherapy. Group Dyn-Theor Res (2000) 4: 115– 134. doi: org/10.1037/1089-2699.4.1.115

35. Joyce AS, MacNair-Semands R, Tasca GA, Ogrodniczuk JS. Factor structure and validity of the Therapeutic Factors Inventory–Short Form. Group Dyn (2011) 15: 201–219. doi: org/10.1037/a0024677

36. Davies DR, Burlingame GM, Jennifer J, Gleave RL, Barlow SH. The Effects of a Feedback Intervention on Group Process and Outcome. Group Dyn-Theor Res (2008) 12: 141–154. doi: 10.1037/1089-2699.12.2.141

36. Brabender V. On the mechanisms and effects of feedback in group psychotherapy. J Contemp Psychother (2006) 36: 121–128. doi: org/10.1007/BF02729055

